# The effects of left vagus nerve block on postoperative nausea and vomiting in patients undergoing thoracic or abdominal laparoscopic surgery: A randomized controlled trial

**DOI:** 10.1101/2024.11.08.24316971

**Authors:** Chen Chen, Zhongyu Yang, Qi Zheng, Yanghao Ren, Tianyu Yang, Xinyue Zhen, Liang Ding, Bingqian Fan, Tianhai Wang, Hongyan Dai

## Abstract

**Purpose:** To explore the influence of preoperative vagus nerve block (VNB) on postoperative nausea and vomiting (PONV) in patients undergoing thoracic or abdominal laparoscopic surgery.

**Methods:** Patients were randomly divided into the VNB group (V group, n=60) and the control group (C group, n=60). The V group received left VNB before anesthesia induction, while the C group did not receive any intervention. The incidence and severity of PONV within one day after surgery were compared between the two groups to evaluate the effect of left VNB on PONV in patients.

**Results:** The incidence of PONV in the V group (25%) was significantly lower than that in the C group (60%) (χ^2^ = 15.038, *P* < 0.001). The incidences of mild and severe PONV in the V group were 16.67% and 8.33%, respectively, while those in the C group were 36.67% and 23.33%, respectively. The differences between the two groups were significant (mild: χ^2^ = 6.136, *P* = 0.013; severe: χ^2^ = 5.065, *P* = 0.024).

**Conclusion:** Left VNB can reduce the incidence and severity of PONV caused by thoracic or abdominal laparoscopic surgery.

## Introduction

Postoperative nausea and vomiting (PONV) is a common complication following surgery. PONV not only prolongs the hospital stay of patients, increases hospitalization costs, and reduces patient satisfaction and quality of life, but also is one of the problems that plague anesthesiologists and surgeons [1–3].

Some studies have shown that the vagus nerve mediates the communication between the gut and brain, which is a key pathway in the occurrence of PONV [4–5]. Scholar Xie *et al*. established a novel paradigm using a mouse model to study the “nausea-vomiting” response and found that vomiting depends on the “gut-to-brain axis,” with the vagus nerve playing an important role [6]. Additionally, an observational cohort study by Li *et al*. indicated that vagus nerve resection during esophagectomy or gastrectomy reduced the incidence of PONV from 28.7% to 11.9% compared to procedures without vagus nerve resection [7]. Although this result suggests that blocking the vagus nerve that controls the gastrointestinal tract may inhibit the occurrence of PONV, the high technical demands on the surgeon, significant visceral trauma, and potential damage to surrounding normal tissue structures limit its clinical application. With the development of ultrasound technology, vagus nerve block (VNB) through the neck has made the injection process visualized, ensuring the safety of the procedure. Additionally, the nerve is surrounded by the carotid sheath, which limits the spread of local anesthetics. Therefore, the dose of local anesthetic required to block this nerve is relatively small, and the block duration is long, which has begun to be applied clinically [8–9].

Therefore, this study aims to explore the effect of left VNB on PONV in patients undergoing thoracic or abdominal laparoscopic surgery, to provide a new choice for the multimodal prevention and treatment strategy of PONV.

## Methods

### Study design and ethics

This study adheres to the Declaration of Helsinki, as revised in 2013 for human trials, and follows the applicable CONSORT guidelines. The study was registered on March 18, 2021, in the Chinese Clinical Trial Registry (Registration No: ChiCTR210004447 0), which is accessible at https://www.chictr.org.cn/searchproj.html. Additionally, on September 20, 2021, the Ethics Committee of the Affiliated Cancer Hospital of Xinjiang Medical University (Chair: Professor Xiu Hua Zhang) granted ethical approval for th is study (Ethics No: K-2021053). The recruitment period of this study was from September 21, 2021 to October 30, 2022. Written informed consent was obtained from patients or their families before anesthesia and nerve block.

### Setting

The trial will be conducted in the Anesthesia and Perioperative Medicine Center of the Affiliated Cancer Hospital of Xinjiang Medical University in Urumqi, Xinjiang, China.

### Patients

A total of 130 patients scheduled for elective thoracic or abdominal laparoscopic surgery were included. A researcher (XZ) who was unaware of the group assignments learned about the patient’s medical histories one day before surgery and signed the informed consent forms for anesthesia and nerve block with the families or patients. The inclusion criteria were: (1) ASA class I-III; (2) aged 25-65 years; (3) no history of PONV; (4) patients undergoing elective thoracic or abdominal laparoscopic surgery. The exclusion criteria were: (1) surgery duration less than 1 hour; (2) intraoperative blood loss over 800 ml; (3) allergy to medications used in the study; (4) neck or systemic infection; (5) neck deformity or large tumors; (6) hoarseness or deaf-muteness; (7) refusal to participate before nerve block.

### Randomization and blinding

A researcher (CC) numbered the patients according to their order of surgery and used randomization software (https://www.randomizer.org) to randomly assign the patients to the VNB group (V group) or the control group (C group) in a 1:1 ratio. For each patient, a sealed opaque envelope was prepared. Before the surgery, a researcher (ZY) opened the envelope in the pre-anesthesia room outside the operating room and decided whether to perform VNB. Both groups of patients were handed over to an anesthesiologist (TY), who administered general anesthesia according to the same procedure. The anesthesiologists (excluding CC and ZY), surgeons, nurses, post-anesthetic care unit (PACU), and outcome assessors were all unaware of the group assignments.

### Perioperative management and intervention

Patients did not receive preoperative medications on the day of surgery or the night before. Patients first entered the pre-anesthesia room outside the operating room, where they underwent standard monitoring procedures. A central venous access was established, and peripheral arterial puncture and catheterization were performed to monitor the patient’s mean arterial pressure (MAP). The patient was connected to a monitor to detect electrocardiogram (ECG), heart rate (HR), pulse oxygen saturation (SpO2), and other indicators.

The V group received ultrasound-guided left VNB in the pre-anesthesia room before anesthesia induction. The patient’s head was turned to the right. The anesthesiologist (ZY) used a high-frequency linear ultrasound probe to perform short-axis scanning at the level of the fourth cervical vertebra transverse process. The internal jugular vein and carotid artery were found. The carotid artery, internal jugular vein, and vagus nerve are all surrounded by the carotid sheath. A round structure with stronger peripheral echoes and weaker central echoes can be detected posterior to the internal jugular vein and lateral to the carotid artery. This structure is the vagus nerve. The needle tip was inserted next to the vagus nerve in the carotid sheath. 0.5 ml of normal saline was injected to determine the needle tip position. After confirming that the needle tip position was correct, 3 ml of 0.5% ropivacaine was injected. During the injection process, attention should be paid to the direction of local anesthetic diffusion. Ideally, the diffusion should be towards the caudal end of the patient. If the drug diffuses towards the cephalic end, the upper part of the puncture site can be gently pressed to prevent the drug from diffusing towards the cephalic end to prevent excessive block level and affecting other nerves. A successful block was indicated by hoarseness within 2-5 minutes. The C group did not undergo VNB, but all other procedures were the same as those for the V group.

Fifteen minutes after the block was completed, the patient was pushed into the operating room. All patients were anesthetized and induced by the same anesthesiologist (TY). Anesthesia induction included midazolam (0.05 mg/kg), propofol (1.5 mg/kg), sufentanil (0.3 μg/kg), and rocuronium (0.6 mg/kg), followed by tracheal intubation. Mechanical ventilation was used for both groups, with a respiratory rate of 10-12 breaths/min, tidal volume of 6-8 ml/kg, and an I: E ratio of 1:2. Anesthesia was maintained with continuous infusions of propofol (2-4 mg/kg/h), remifentanil (0.5 μg/kg/h), and rocuronium (0.5 mg/kg/h). Additional sufentanil (0.15 μg/kg) was administered at the end of surgery. All patients were given patient-controlled intravenous analgesia pumps after surgery. The drugs for patient-controlled analgesia were all 4 mg of butorphanol tartrate and 100 μg of sufentanil diluted to 100 ml with normal saline, with a flow rate of 2 ml/h.

After extubation, the patient was transferred to the PACU, where the same anesthesiologist (YR) was responsible for monitoring for at least 30 minutes until the modified Aldrete score reached more than 9 points as the discharge standard from the PACU. After the patient was sent back to the ward, the same clinician (QZ) was responsible. Whether in the PACU or the ward, no antiemetic drugs were given to the patient before the occurrence of postoperative nausea and vomiting. In addition, patients and their families were informed in advance to pay attention to the occurrence of nausea and vomiting within 24 hours after surgery, and the patient was arranged in a quiet ward to keep them away from factors that may induce nausea and vomiting.

### Observation indicators

The collection of data was uniformly carried out by trained researchers (BF and LD). Preoperative general data of the two groups of patients were recorded: age, gender, weight, body mass index (BMI), American Society of Anesthesiologists (ASA) classification, and smoking history. Postoperative data recorded included the site of surgery, type of surgery, time of surgery, and the dosage of sufentanil used intraoperatively.

The primary outcomes were the incidence and severity of PONV on the first postoperative day. PONV is defined as the occurrence of nausea and vomiting within 24 hours after surgery. Nausea is assessed based on the patient’s subjective feeling of nausea or the urge to vomit. Both retching and vomiting are recorded as vomiting episodes. Retching is defined as the absence of gastric content expulsion, while any expulsion of gastric contents is recorded as vomiting. The severity is classified as mild (< 3 episodes/day) and severe (≥3 episodes/day). Patients with only retching symptoms are recorded as having mild vomiting [10].

The secondary outcomes included: (1) the duration of hoarseness in the V group. (2) HR and MAP at T0 (10 minutes before nerve block), T1 (5 minutes before nerve block), T2 (during nerve block), T3 (5 minutes after nerve block), T4 (10 minutes after nerve block), and T5 (15 minutes after nerve block).

### Sample size calculation and statistical analysis

We calculated the number of enrolled patients based on the incidence of PONV using PASS2021 software. Firstly, we assessed that the incidence of PONV in 34 patients undergoing laparoscopic surgery was 50% (these 34 patients were not included in the subsequent trial). After patients received the intervention, the reduction in the incidence of PONV showed differences in different studies. The recorded differences were 26% [11], 28% [12], and 37.7% [13]. Therefore, We believe that a 30% reduction in the incidence of PONV is clinically significant. We hypothesized that after implementing left VNB, the incidence of PONV would be 20%, with an α level of 0.05, power (1-β) of 0.90, and an attrition rate of 20%, resulting in a required sample size of 65 patients per group.

In this study, IBM SPSS Statistics 26 software was used for data processing. Continuous variables that conformed to a normal distribution were expressed as mean ±standard deviation and compared between groups using the two independent samples *t*-test. Continuous variables that did not conform to a normal distribution were expressed as Median (Q1, Q3) and compared between groups using the Mann-Whitney U test. Categorical variables were described as number (percentages) and compared between groups using the chi-square test. A *P*-value less than 0.05 was considered statistically significant.

## Results

### Patients’ characteristic

Between September 2021 and October 2022, a total of 130 patients were enrolled, of whom 2 were excluded based on exclusion criteria and 4 refused to participate. A total of 124 patients were recruited and randomly assigned to one of two groups, with 62 patients in each group. One patient in the V group refused to accept the block before the procedure. Three patients were later excluded due to failure to complete follow-up. A total of 120 patients completed the study, including 60 in the V group and 60 in the C group (**Fig. 1**). There were no significant differences in patient characteristics such as age, weight, BMI, gender, ASA classification, smoking status, site of surgery, type of surgery, time of surgery, and intraoperative sufentanil dosage between the two groups (**Table 1**).

**Fig.1.**
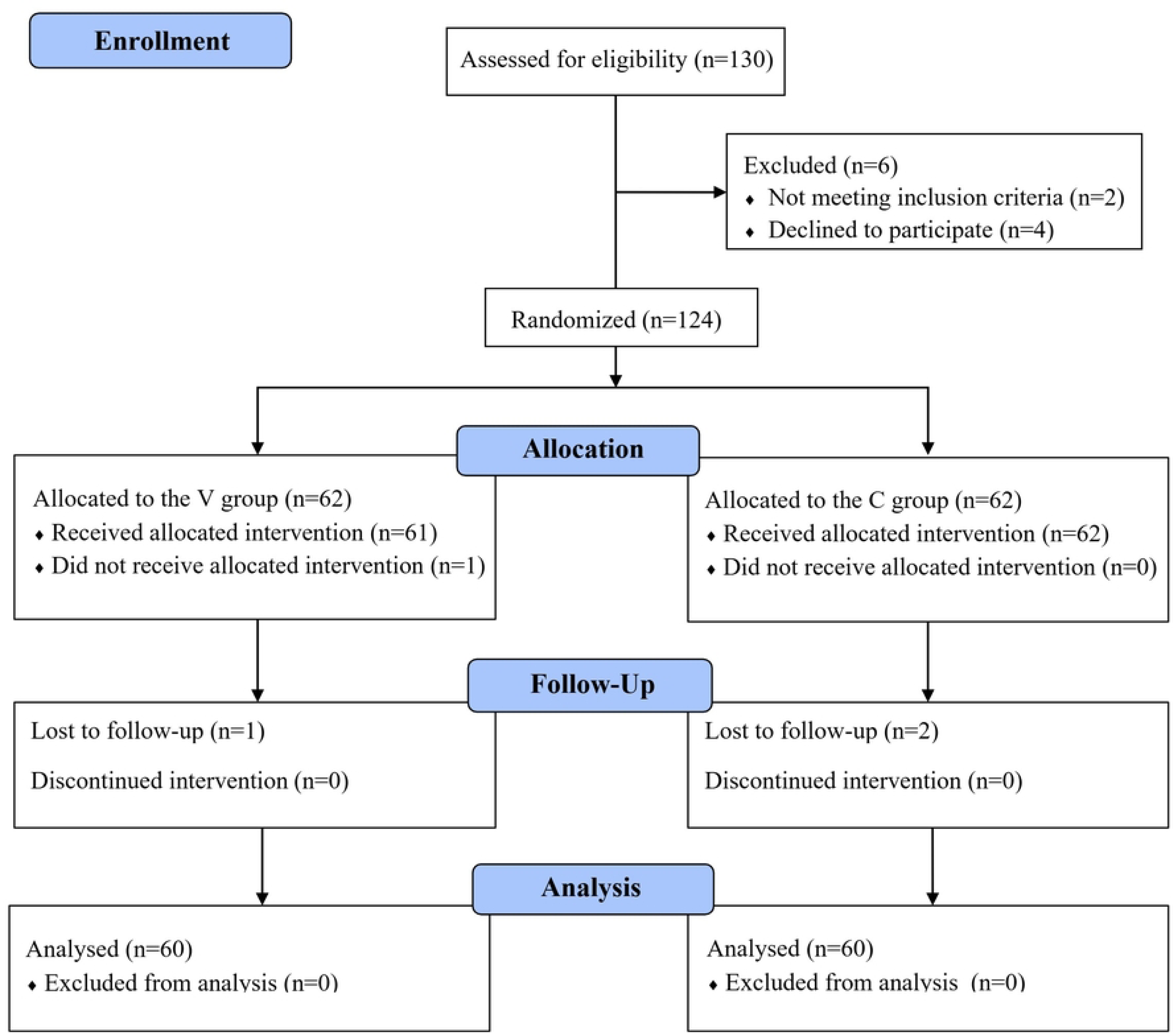
Trial flow chart.

**Table 1.**
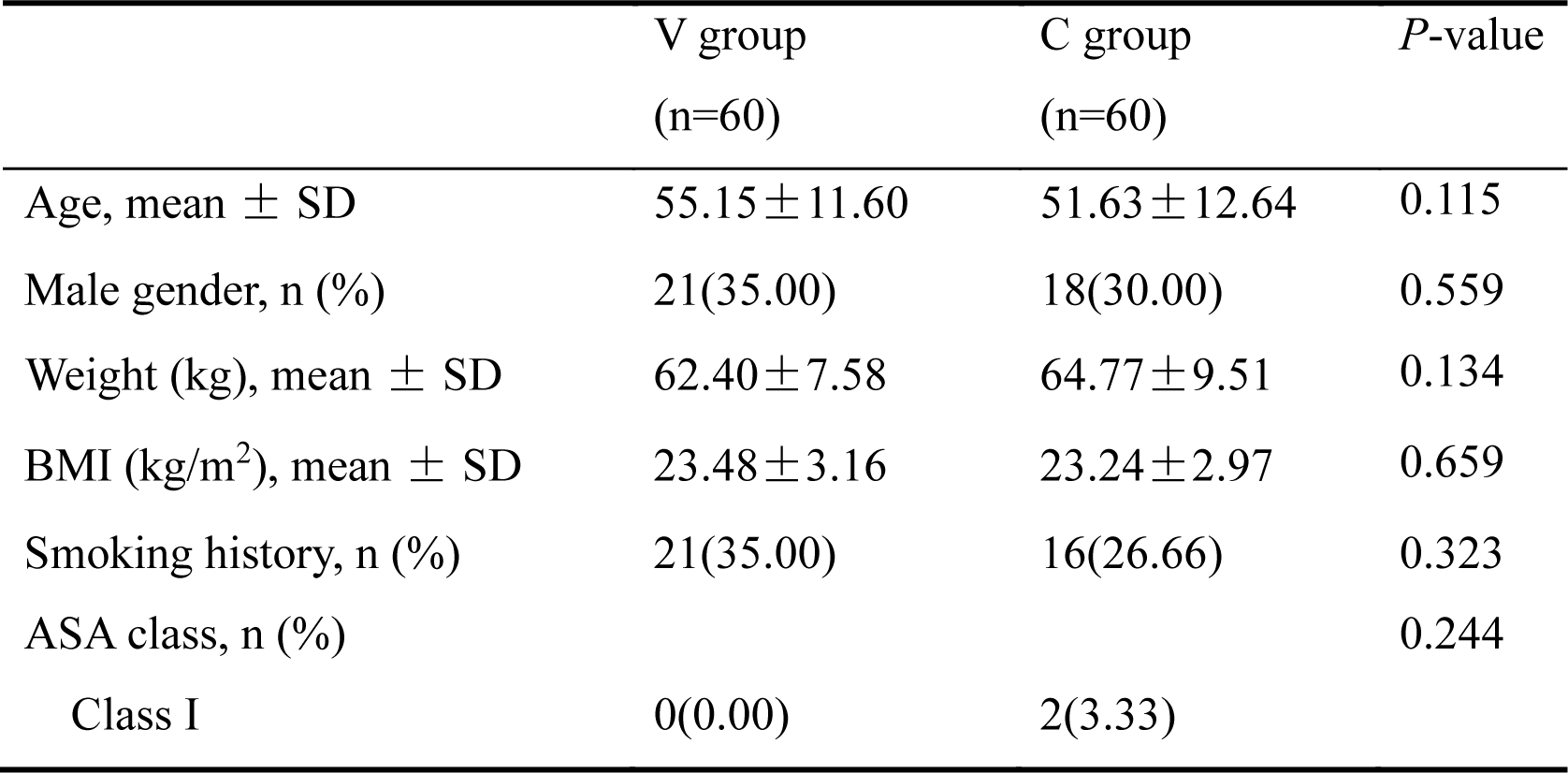
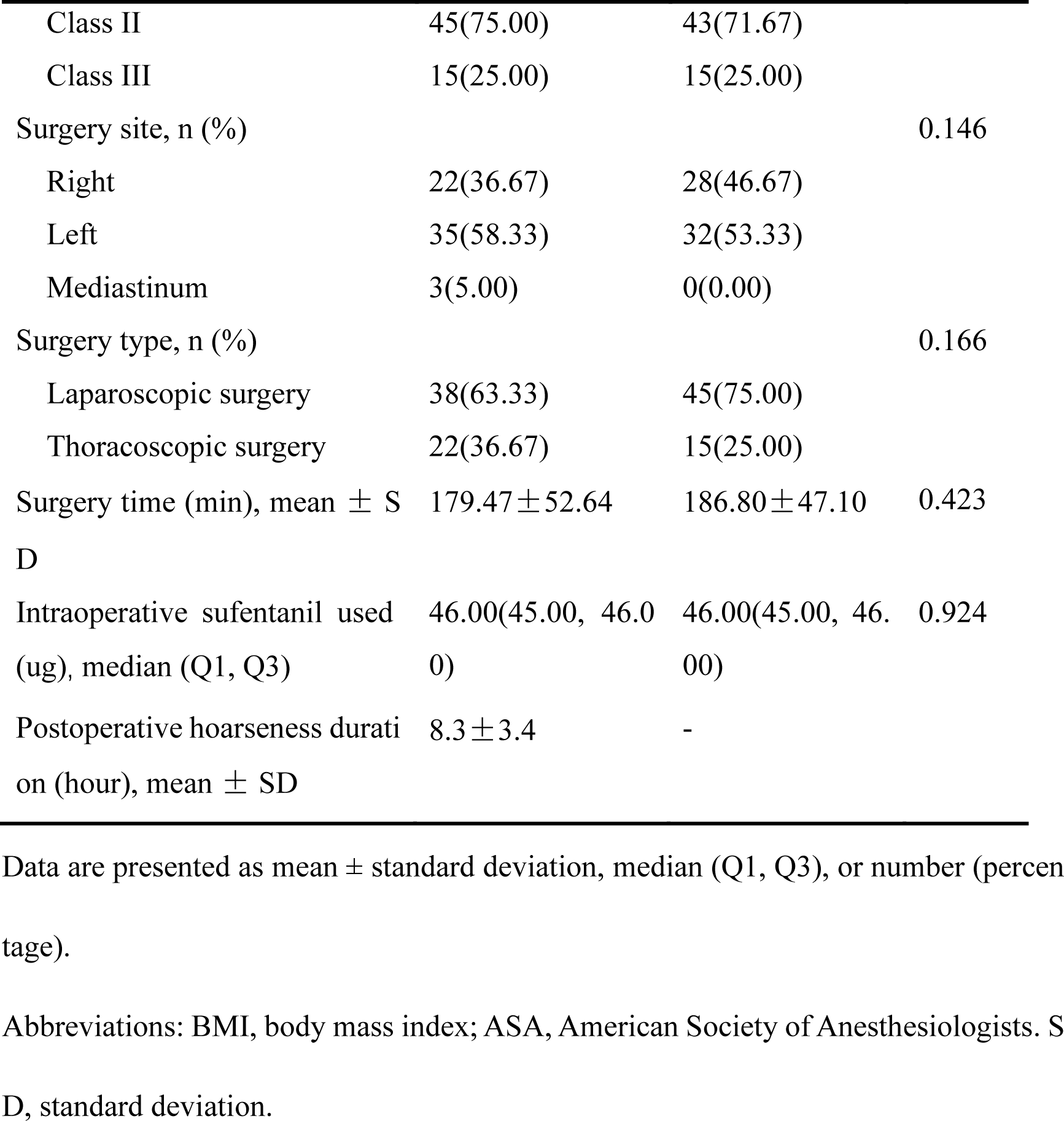
Demographic and surgical characters.

### Primary outcomes

The incidence of PONV in the V group (25%) was significantly lower than that in the C group (60%) (χ^2^ = 15.038, *P* < 0.001, **Table 2**). The incidences of mild and severe PONV in the V group were 16.67% and 8.33%, respectively, while in the C group, they were 36.67% and 23.33%, respectively. The differences between the two groups were statistically significant (mild: χ^2^ = 6.136, *P* = 0.013; severe: χ^2^ = 5.065, *P* = 0.024, **Table 2**).

**Table 2.** Incidence and severity of PONV between the two groups.

| | V group<br>(n=60) | C group<br>(n=60) | $\chi^2$ | *P*-value |
| --- | --- | --- | --- | --- |
| Incidence of PONV, n (%) | 15(25.00) | 36(60.00) | 15.038 | <0.001 |
| Severity of PONV, n (%) |  |  |  |  |
| Mild | 10(16.67) | 22(36.67) | 6.136 | 0.013 |
| Severe | 5(8.33) | 14(23.33) | 5.065 | 0.024 |

Data are presented as numbers (percentages).
Abbreviations: PONV, postoperation nausea and vomiting.

### Secondary outcomes

There were no significant differences in HR and MAP between the two groups at the same time points and between T1-T5 and T0 within each group (**Fig. 2**). The duration of hoarseness in the V group was 8.3 ± 3.4 hours (**Table 1**).

**Fig.2.**
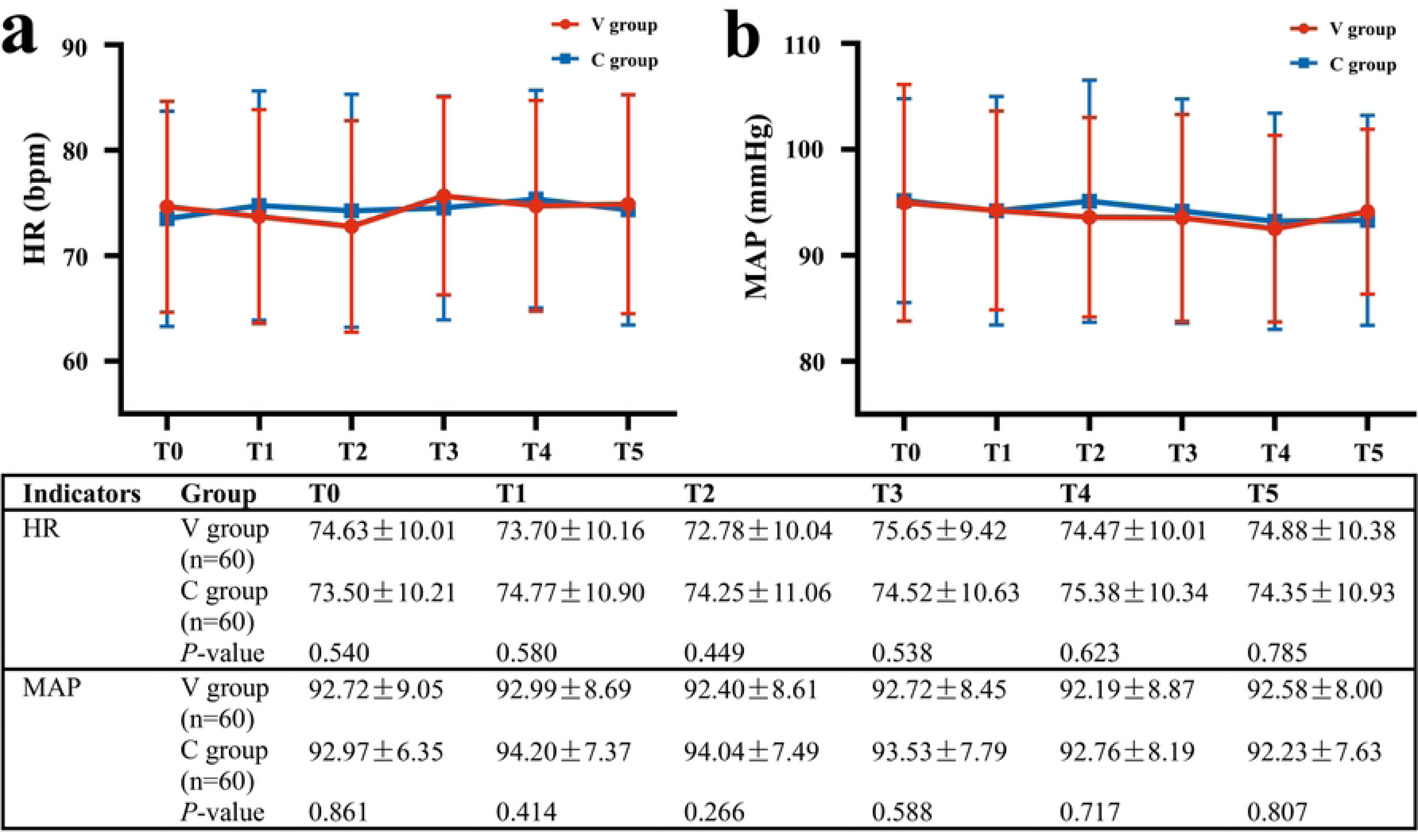
Comparison of HR and MAP at the same time points between the two groups and before and after nerve block within each group. T0: 10 minutes before nerve block; T1: 5 minutes before nerve block; T2: during nerve block; T3: 5 minutes after nerve block; T4: 10 minutes after nerve block; T5: 15 minutes after nerve block. MAP: Mean arterial pressure; HR: Heart rate.

## Discussion

In this study, we tracked the incidence of PONV within postoperative 24 hours. The results showed that the V group had a significantly lower incidence of PONV than the C group by 35% (**Table 2**). Additionally, comparing the severity of PONV between the two groups, the V group had fewer cases of mild and severe PONV, indicating that VNB can reduce the incidence and severity of PONV.

The vagus nerve, which is widely distributed in the gastrointestinal tract, transmits signals to the vomiting centers in the medulla’s lateral reticular formation upon stimulation, enhancing the excitability of these centers [14]. These centers send instructions through the vagus nerve, sympathetic nerve, and phrenic nerve to the stomach and other related organs, triggering the vomiting response [15]. Related studies suggest that the gut-vagus-brain reflex pathway, a typical emetic neural conduction pathway, plays a key role in PONV occurrence [6]. Studies have shown that gastrointestinal-related surgeries or traumas can cause the release of 5-HT_3_ from enterochromaffin cells, leading to the activation of 5-HT_3_ receptors at the vagus nerve afferent nerve endings, which in turn induces vagus nerve reflexes, nausea, and vomiting [16–17]. Similarly, anesthetics can enhance vagal afferent excitation by activating substance P and 5-HT_3_ receptors [14]. Additionally, patients undergoing thoracic or abdominal laparoscopic surgery typically have higher intrathoracic and intra-abdominal pressures, and hypercapnia can activate related vagal reflexes, inducing nausea and vomiting [18–19]. The release of inflammatory mediators and cytokines caused by surgery mainly occurs within 24 hours after surgery [20–21], which suggests that the vagus nerve of patients may be in a state of high stimulation within 24 hours after surgery. We observed the duration of postoperative hoarseness in the V group (**Table 1**) and speculated that the average duration of left VNB was about 8 hours. Based on this, we hypothesize that left VNB can reduce the conduction of signals to the vomiting centers such as the dorsolateral margin of the lateral reticular formation of the medulla within 8 hours after surgery, thereby maintaining the excitability of the vomiting center at a lower level. The vomiting signals sent by the vomiting center to the efferent nerves are reduced, ultimately reducing the incidence and severity of PONV in patients. Therefore, we conclude that the left vagus nerve pathway in the neck may be one of the key factors in the occurrence of PONV. Blocking this neural conduction pathway can effectively reduce the incidence and severity of PONV.

In this study, we observed the effects of left VNB on the HR and MAP of patients at six-time points from T0 to T5. The results showed that there was no significant difference in HR and MAP before and after the block within the same group and at the same time points between the two groups (**Fig. 2**), indicating that left VNB has limited effects on HR and MAP. Considering the reasons: The heart is regulated by bilateral vagus nerves. The right vagus nerve dominates the function of the sinoatrial node and is more closely related to the atrium. The left vagus nerve is mainly related to the ventricle, and the vagus nerve innervation of the ventricle is not as dense as that of the atrium [22]. Therefore, left VNB generally does not have a significant impact on the heart.

This study has the following limitations: First, as a single-center study, the sample size is limited. Broader research and clinical validation are needed to confirm the universality of these findings. Second, this study only focused on the incidence and severity of PONV within the first day after surgery. No in-depth study was conducted on the situation for a longer time after surgery. In the future, the long-term effects of left VNB on PONV still need to be considered.

In summary, preoperative left VNB can prevent the incidence and severity of PONV caused by thoracic or abdominal surgery. Additionally, left VNB has no significant impact on hemodynamics (HR and MAP). Left VNB can be considered as a new option for preventing the occurrence of PONV.

## Author contributions

***Chen Chen:*** 333 data curation, formal analysis, methodology, project administration, software, validation, visualization, writing - original draft, writing - review & editing. ***Zhongyu Yang:*** performing VNB, methodology, writing original draft, review & editing. ***Qi Zheng:*** patient management, investigation, methodology. ***Yanghao Ren:*** patient management, investigation, methodology. ***Tianyu Yang:*** patient management, investigation, methodology. ***Xinyue Zhen:*** investigation, methodology. ***Liang Ding:*** data collection. ***Bingqian Fan:*** data collection. ***Tianhai Wang:*** conceptualization, formal analysis, funding acquisition, methodology, project administration, resources, supervision, validation, writing - review & editing; ***Hongyan Dai:*** conceptualization, formal analysis, methodology, project administration, resources, softw are, supervision, validation.

## Data Availability

All relevant data are within the manuscript and its Supporting Information files.

## Acknowledgments

None.

## Disclosures

The authors report no conflicts of interest.

## Funding statement

None.

## Notes

### Competing Interest Statement

The authors have declared no competing interest.

### Clinical Trial

Trial was prospectively registered in the Chinese Clinical Trial Registry (number: ChiCTR2100044470).

### Clinical Protocols

https://www.chictr.org.cn/searchproj.html

### Funding Statement

The author(s) received no specific funding for this work.

### Author Declarations

On September 20, 2021, the Ethics Committee of the Affiliated Cancer Hospital of Xinjiang Medical University (Chair: Professor Xiu Hua Zhang) granted ethical approval for this study (Ethics No: K-2021053).

